# 7 Tesla MRI links poorer cognitive function to higher perivascular space burden in neuroPASC

**DOI:** 10.64898/2026.05.01.26352247

**Authors:** Mackenzie T. Herb, Jacqueline H. Becker, David O’Connor, Matthew R. Perez, Sera Saju, Yijuan Zhu, Gaurav Verma, Nathalie Jette, Bradley N. Delman, Priti Balchandani, Alan C. Seifert

**Author notes:** **Correspondence:** Mackenzie T. Herb, One Gustave L. Levy Pl., Box 1234, New York, NY 10029.

## Abstract

**Purpose:** Post-acute sequalae of SARS-CoV-2 (PASC) are associated with persistent neurological symptoms (neuroPASC). Perivascular spaces (PVS) in the brain may enlarge in the context of inflammation and vascular dysfunction, reflecting impaired glymphatic clearance, and have been linked to cognitive decline. SARS-CoV-2 may disrupt the blood-brain barrier and impair glymphatic function, contributing to PVS burden. This study used 7 Tesla MRI to segment and quantify PVS in neuroPASC participants and uninfected comparators and examined associations with cognitive performance.

**Methods:** Adult participants (36 neuroPASC (44.3 ± 12.7 years) and 33 comparators (38.4 ± 13.0 years)) underwent a 7 Tesla MRI scan. White matter masks of the whole brain and four lobes were segmented, and semi-automated segmentation was used to quantify PVS count and volume. All participants completed cognitive testing including Trails A and B sequencing tasks; neuroPASC participants also self-reported brain fog, fatigue, anxiety, and depression. PVS count, PVS volume, and total white matter volume (WMV) between groups were compared and associations between PVS metrics and cognitive function were assessed controlling for age, sex, and intracranial volume and corrected for multiple comparisons.

**Results:** Among neuroPASC participants, those reporting anxiety (*p* =0.009) and depression (*p* =0.01) had higher WMV than those without. Greater PVS burden was associated with worse cognitive performance in PASC, particularly processing speed (Trails A) and executive function (Trails B). Specifically, processing speed was negatively associated with whole-brain PVS count (*p-FDR* = 0.008, *R*^*2*^ = 0.27), frontal PVS count (*p-FDR* = 0.03, *R*^*2*^ = 0.25), and frontal PVS volume (*p-FDR* = 0.04, *R*^*2*^ = 0.23). Trails B was also negatively associated with whole-brain PVS count (*p-FDR* = 0.005, *R*^*2*^ = 0.26). In comparators, higher PVS burden (volume and count) across multiple lobes was associated with worse semantic fluency (Animal Naming). There were no other significant associations between PVS measures and neuropsychiatric tests among participants within any of the subgroups to report.

**Conclusion:** Although group-level differences in PVS were not observed, PVS burden was meaningfully negatively associated with cognitive performance in neuroPASC, with the strongest effects in frontal regions. These findings suggest that microvascular and glymphatic alterations may contribute to the characteristic processing speed and executive dysfunction seen in neuroPASC. Elevated WMV in those with anxiety and depression may reflect heightened inflammatory vulnerability. PVS may serve as a sensitive imaging marker of glymphatic dysfunction and neuroinflammation in neuroPASC, offering insight into the mechanisms underlying cognitive impairment and potential intervention targets.

## Introduction

SARS-CoV-2 infection has been associated with a wide range of neurological and neuropsychiatric sequelae, including cognitive impairment, fatigue, depression, and anxiety, which in many individuals persist well beyond the acute phase.^1^ The presence of persistent symptoms, collectively referred to as post-acute-sequelae of SARS-CoV-2 (PASC), affect a substantial proportion of COVID-19 survivors and represent a major public health burden.^2^ Among the most common and debilitating complaints are cognitive difficulties, often described as “brain fog,” which encompass deficits in processing speed, attention, executive function, and verbal memory.^3,4^ Despite the clinical significance of these symptoms, the underlying neuropathological mechanisms remain poorly understood.

Perivascular spaces (PVS), also known as Virchow-Robin spaces, are fluid-filled compartments that surround penetrating blood vessels as they course through the brain parenchyma.^5^ Under normal physiological conditions, PVS serve as conduits for cerebrospinal fluid (CSF) transport and interstitial fluid drainage, playing a critical role in the glymphatic system.^6-9^ The glymphatic system facilitates the exchange of CSF and interstitial fluid, enabling the removal of metabolic waste products, including beta amyloid and tau proteins.^10-12^

Increased PVS burden, visible on MRI, is increasingly recognized as a marker of impaired glymphatic function and has been implicated in neuroinflammatory processes, blood-brain barrier (BBB) dysfunction, and cerebral small vessel disease.^5,13,14^ When PVS become dilated or enlarged, they may reflect obstruction of normal perivascular drainage, accumulation of inflammatory cells, or downstream consequences of vascular stiffening and reduced arterial pulsatility.^15^ One proposed mechanism is that PVS enlargement may be a structural consequence of underlying pathology and also reflects glymphatic clearance which may contribute to the accumulation of neurotoxic waste products and subsequent neuronal injury.^16^ More specifically, white matter PVS accumulation and enlargement is associated with increased microglial activation and neuroinflammation, which suggests that inflammatory processes may drive PVS burden.^17^ Neuroinflammation can cause abnormal neural network activity through multiple different mechanisms leading to cognitive and executive function impairment. ^18,19^

A growing body of evidence links increased PVS burden to cognitive impairment across diverse clinical populations.^20^ Meta-analytic data indicate that increased PVS burden is significantly associated with cognitive impairment, particularly in executive function, processing speed, semantic memory, and global cognitive function.^21-23^ These findings suggest that higher PVS burden is a clinically meaningful marker of neurovascular or inflammatory contributions to cognitive dysfunction.

SARS-CoV-2 has been shown to disrupt the BBB, trigger perivascular inflammation, and potentially impair glymphatic clearance, all of which could contribute to increased PVS burden in COVID-19 survivors.^24,25^ Recent neuroimaging studies have demonstrated glymphatic dysfunction in PASC participants using diffusion-based MRI metrics, and associations between glymphatic impairment and neurocognitive symptoms have been observed.^26,27^ Our group previously demonstrated in a pilot study that participants with COVID-19 exhibited increased PVS burden and white matter volume compared to healthy controls with no history of SARS-CoV-2 infection using 7 Tesla (7T) MRI.^28^ However, that study did not examine the relationship between PVS burden and neurocognitive function.

Ultra-high field 7T MRI offers substantial advantages for PVS quantification over conventional clinical field strengths, providing superior signal-to-noise and contrast-to-noise ratios that enable the detection and characterization of smaller PVS that would otherwise be undetectable at 1.5T or 3T.^29-31^ This improved resolution is particularly valuable for studying PVS in the context of disease, as it allows for more precise segmentation and quantification of PVS across brain regions.^32,33^

The present study leverages 7T MRI to segment and quantify PVS in a cohort of participants with persistent neurological symptoms following SARS-CoV2 infection (neuroPASC) compared to comparators without persistent neurological symptoms. We examined group-level differences in PVS burden between participants and comparators as well as within symptom-defined subgroups of the neuroPASC cohort (brain fog, fatigue, depression, and anxiety). Additionally, we investigated associations between PVS burden and performance on neuropsychological measures of processing speed, executive function, verbal memory, and working memory. We hypothesized that greater PVS burden, particularly in frontal regions, would be associated with worse cognitive performance among participants with neuroPASC, reflecting the impact of neuroinflammation and impaired glymphatic function on cognitive efficiency.

## Methods

### Participants

We recruited 51 neuroPASC participants and 43 comparators through the Mount Sinai health system (Table 1). All participants were enrolled between 2021-2025 and were included in the neurological symptom-positive group if they had a self-reported or documented history of SARS-CoV-2 infection, and reported at least one persistent neurological symptom or diagnosis (e.g., anosmia, altered taste/smell, brain fog, fatigue, stroke, encephalopathy, delirium, memory impairment) that lasted for at least 3 months after infection. Participants with a pre-COVID history of major neurological or neuropsychiatric conditions or those ineligible for MRI (e.g., due to implants or pregnancy) were excluded. Comparator participants had the same inclusion/exclusion criteria except had no reported persistent symptoms of COVID-19. Written informed consent was obtained from all participants prior to scanning. The Mount Sinai Health System Institutional Review Board approved this study. Participants’ self-reported symptoms were collected from all COVID-19 participants at the time of visit including presence of brain fog, anxiety, fatigue, and depression (Table 1). Sixteen neuroPASC participants and ten comparators were excluded due to excessive motion during MRI scan.

**Table 1.** Demographics.

|  | neuroPASC | Comparators | p-value |
| --- | --- | --- | --- |
| N | 36 | 33 | - |
| Age [mean ± SD] | 44.3 ± 12.7 | 38.4 ± 13.0 | 0.06 |
| Sex [Number of females/males] | 22/14 | 18/15 | 0.80 |
| **Symptom** | **N with symptom**<br>**[number of females/males]** | **N without symptom**<br>**[number of females/males]** |  |
| Brain Fog | 27 [16 F/ 11M] | 9 [6 F/ 3 M] |  |
| Anxiety | 22 [12 F/ 10 M] | 14 [10 F/ 4 M] |  |
| Depression | 18 [9 F/ 9 M] | 18 [13 F/ 5 M] |  |
| Fatigue | 31 [19 F/ 12M] | 5 [3 F/ 2M] |  |

SD: standard deviation

### Imaging acquisition and processing

All subjects underwent preoperative scanning on a Siemens Magnetom 7 T scanner which included T1-weighted MP2RAGE (TR: 6000 ms, TE: 3.62 ms, TI (TI2): 1050 (3000) ms, FOV: 168 mm × 244 mm, number of slices: 240, resolution: 0.7 mm isotropic), and an axial T2TSE (minimum TR = 9,000.0 ms, TE = 59 ms, flip angle = 158°, FOV = 168 × 200 mm2, matrix = 512 × 432, in-plane resolution 0.2 × 0.2 mm2, slice thickness = 2 mm, slice gap = 0.6 mm, slice number = 56, BW = 279 Hz/pixel, minimum time = 6:21 min).

T1 image data were preprocessed using FreeSurfer 7.4.1 including skull stripping, motion correction, and normalization.^34,35^ Outputs from FreeSurfer reconstruction were used to segment white matter masks of the whole brain and each lobe. All patient T2TSE images were visually inspected for quality, and participants with excessive motion were excluded (neuroPASC n = 16, comparators n = 10) from further processing as motion artifacts impede PVS segmentation. A semiautomated segmentation tool called perivascular space semi-automated segmentation (PVSSAS) ^28,36^ was implemented to segment and quantify white matter PVS, blinded to patient status, from whole brain and each brain lobe. Total PVS count and volume (voxels) were quantified in all subjects.

### Neuropsychiatric Testing

All subjects underwent neuropsychological testing across several domains, including the Hopkins Verbal Learning Test-Revised (HVLT-R), Trail Making Tests (TMT) Parts A and B, Number Span Forward and Backward (total correct and longest span), and Wide Range Achievement Test-4 (WRAT-4). Raw scores were transformed into z-scores based on published normative data accounting for age, sex, and education. For all tests, lower z-scores indicate worse performance.

### Statistical Analysis

Differences in PVS count and total volume of whole brain and each lobe were calculated using logistic regression controlling for age, sex, and total intracranial volume (ICV) between neuroPASC (n = 36) and comparators (n=33) using MATLAB (2024b). Within the neuroPASC cohort, four additional subgroup analyses were performed for participants reporting symptoms compared to those without including: brain fog, fatigue, depression, and anxiety. Additionally, PVS metrics were corrected for age and sex, and regression analyses were performed for each group (PASC, comparator, and each subgroup) investigating the relationship between neuropsychiatric test z-scores and residual PVS metrics controlling for ICV. Within the subgroup analysis co-incidence of symptoms was controlled for in addition to ICV. Results were corrected for multiple comparisons using false discovery rate (FDR) correction with significance level p < 0.05 and R^2^ values were used to assess affect size between PVS metrics and neuropsychiatric performance. Associations within the neuroPASC subgroup reporting no fatigue (n =5) and no brain fog (n=9) were not investigated due to insufficient statistical power resulting from the sample size. Hypothesis tests were grouped into two a priori defined metrics based on outcome type: PVS volume and PVS count wherein each metric had four regional tests (frontal, parietal, temporal and occipital lobes) and FDR correction was applied separately within each metric. This approach was chosen since PVS count and volume are considered to capture distinct, yet related aspects of PVS burden and therefore treated as separate outcomes.

## Results

### Group-level Analysis

Among neuroPASC participants, total white matter volume was significantly greater among those reporting anxiety (*p*=0.009) and depression (*p*=0.01) compared to those without. There were no significant differences in whole brain PVS count, total PVS volume, regional-lobe PVS count nor volume, and total white matter volume in any of the lobes, between neuroPASC participants and comparators. Within the neuroPASC groups, there were no differences across any PVS metric within each of the subgroups (fatigue v. no-fatigue, brain fog v. no-brain fog, anxiety v. no-anxiety, depression v. no-depression).

### Association with Neuropsychiatric Tests

#### Comparators

Among comparators, across all four lobes, there were negative associations between PVS total volume and semantic fluency. Additionally, PVS count within the temporal, parietal, and occipital lobes was negatively associated with semantic fluency (Table 2).

**Table 2:** Associations between PVS and Semantic Fluency within Comparators.

| Region | PVS count |  | PVS total Volume |  |
| --- | --- | --- | --- | --- |
|  | p-FDR | R<sup>2</sup> | p-FDR | R<sup>2</sup> |
| Whole Brain | 0.78 | 0.02 | 0.09 | 0.15 |
| Frontal | 0.06 | 0.13 | 0.04 | 0.16 |
| Temporal | 0.01 | 0.26 | 0.003 | 0.34 |
| Parietal | 0.02 | 0.20 | 0.01 | 0.22 |
| Occipital | 0.01 | 0.27 | 0.01 | 0.27 |

#### NeuroPASC Participants

Across all neuroPASC participants, there were significant relationships between Trails A and whole brain PVS count. This relationship was also found between Trails A and frontal lobe PVS count and PVS total volume (Figure 1). Additionally, there were significant associations between whole brain PVS count and Trails B performance (Figure 2). No significant associations between PVS measures and neuropsychiatric tests among participants within any of the subgroups survived correction for multiple comparisons.

**Figure 1.**
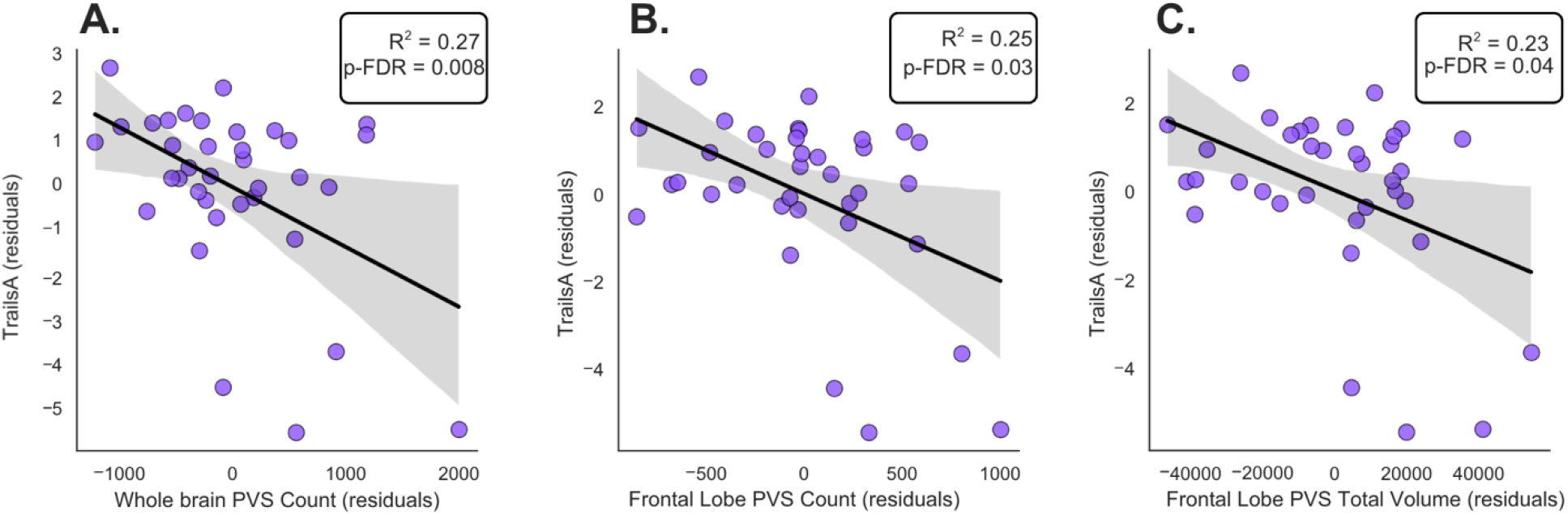
Associations between PVS Metrics and Trails A z-score among all neuroPASC participants. Residual values for Trails A (z-score) and whole brain PVS count (A), frontal lobe PVS count (B), and frontal lobe PVS total volume (C) are shown. Each point represents an individual participant and regression line reflects the fit adjusted for ICV. Statistical significance was determined using FDR correction for multiple comparisons.

**Figure 2.**
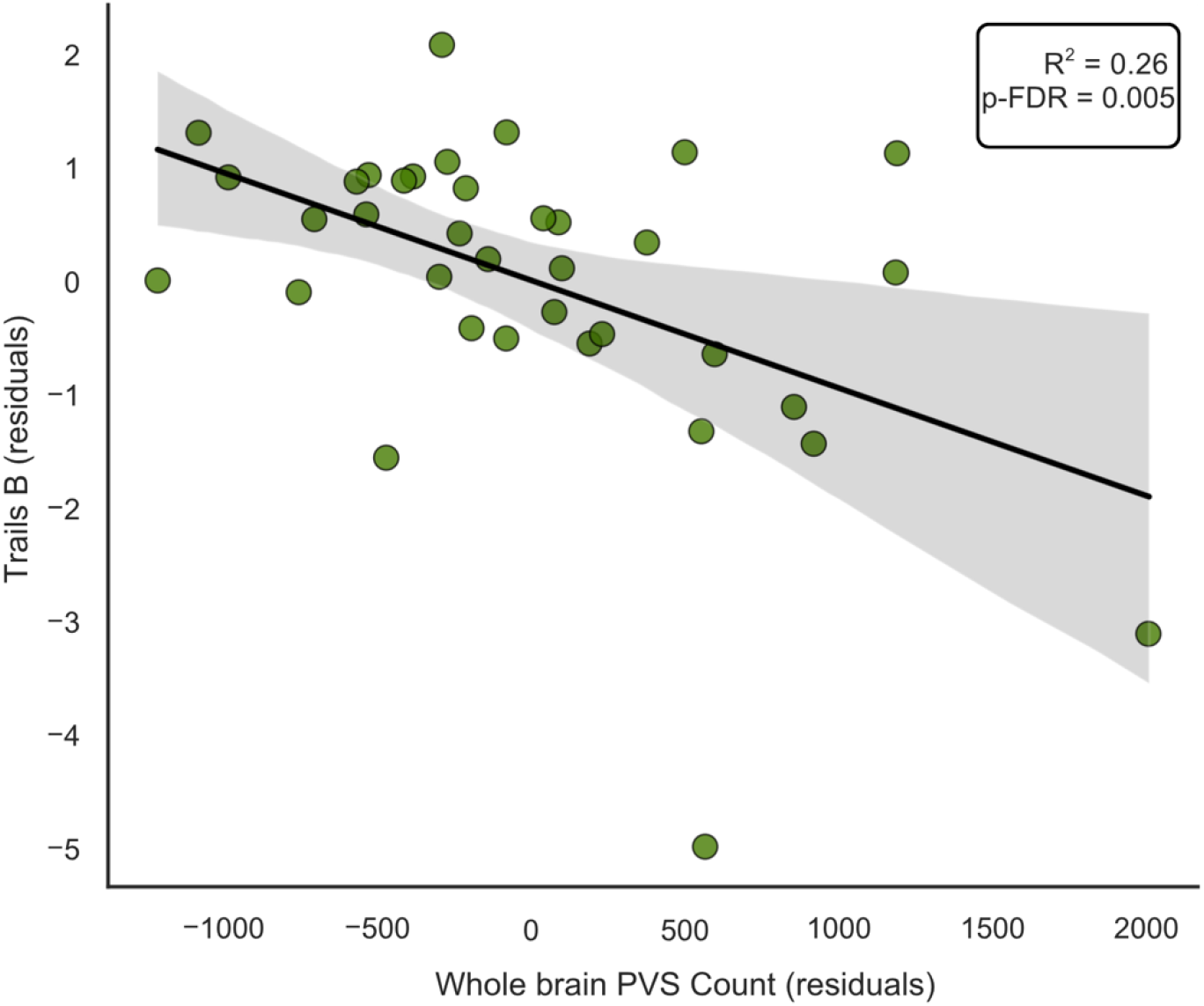
Associations between Whole Brain PVS Count and Trails B z-score among neuroPASC participants. Residual values for Trails B (z-score) and whole brain PVS count shown. Each point represents an individual participant and regression line reflects the fit adjusted for ICV. Statistical significance was determined using FDR correction for multiple comparisons.

## Discussion

This study used ultra-high field 7T MRI to investigate perivascular space burden and its associations with neuropsychological performance in neuroPASC participants. While no significant group-level differences in PVS count or volume were observed between neuroPASC participants and comparators, several clinically meaningful brain-behavior associations emerged. Among neuroPASC participants, greater PVS burden—particularly in the frontal lobe—was associated with slower processing speed and reduced executive function as measured by Trail Making Tests A and B.

The absence of group-level differences in PVS burden between neuroPASC participants and comparators warrants further interpretation. PASC is a clinically heterogeneous condition encompassing diverse symptom profiles, variable severity, spatial heterogeneity, and likely multiple underlying pathophysiological mechanisms.^37^ This heterogeneity may obscure group-wise differences when comparing all neuroPASC participants to comparators, as individual variation in PVS burden may relate more closely to specific symptom patterns or disease mechanisms than to neuroPASC status per se. Furthermore, higher PVS burden may represent a more gradual, cumulative process rather than an acute or sub-acute response to infection, such that cross-sectional comparisons between groups may not capture the full extent of ongoing neurovascular changes.^26,38,39^ It is also possible that PVS serve as a lagging indicator of neurovascular dysfunction: cognitive impairment and neurological symptoms may precede detectable changes in PVS morphology on imaging, with PVS accumulation reflecting a downstream consequence of sustained neuroinflammation rather than an early biomarker.^15,16,23,40^

Despite the lack of group-level differences, the significant associations between PVS metrics and Trail Making Test performance among neuroPASC participants are noteworthy. Trails A primarily indexes processing speed and psychomotor function, while Trails B additionally engages cognitive flexibility and set-shifting, core components of executive function.^41,42^ The negative associations observed between frontal lobe PVS burden and Trails A and B performance suggest that PVS accumulation within frontal white matter may reflect disrupted neural circuits supporting these capacities.^43,44^ This interpretation is consistent with the broader literature linking enlarged PVS to reduced information processing speed in the context of cerebral small vessel disease, and with evidence that frontal PVS burden specifically associated with the disruption of the integrity of white matter tracts subserving executive processes.^45-48^ Theoretically, PVS burden in the frontal lobes could impair local fluid and waste drainage, compromise the microstructural environment of surrounding white matter, and thereby reduce the efficiency of frontal-subcortical circuits that underpin speeded cognitive performance. ^49-52^

In comparators, PVS burden was significantly associated with semantic fluency, with negative associations observed across temporal, parietal, and occipital lobes. This finding aligns with evidence that higher PVS burden is associated with diminished verbal and semantic processing even in the absence of clinical disease, and may reflect age- or vascular-related changes in glymphatic function among otherwise healthy individuals.^21,53^ The divergence in cognitive correlates between comparators (semantic fluency) and neuroPASC participants (processing speed and executive function) may reflect distinct pathological mechanisms: while PVS-related cognitive associations in healthy aging may be driven by gradual neurovascular changes, in neuroPASC participants the pattern may be shaped by post-infectious neuroinflammation preferentially affecting frontal-executive circuitry. ^54,55^

In neuroPASC participants reporting anxiety and depression, increased white matter volume was found compared to those without. In neuroPASC, increased white matter volume could be indicative of edema or increased extracellular free water secondary to neuroinflammation mediated BBB disruption rather than true tissue expansion.^54,56^ Since this difference was only reflected among the neuroPASC participants who reported anxiety or depression and not within the entire cohort, this could be indicative that participants with co-occurrences of neuropsychiatric conditions may be more susceptible to BBB dysfunction mediated by COVID-19 related systemic inflammation along shared pathways compared to individuals without prior SARS-CoV-2 infection. ^28,57,58^ Current literature reports widespread white matter microstructural changes and increased PVS among patients with reported anxiety and depression but typically white matter volume and integrity are reduced. ^59,60^

From a mechanistic perspective, SARS-CoV-2 is known to disrupt the BBB and trigger perivascular inflammation, potentially impairing the glymphatic clearance system.^25,26^ Perivascular inflammation may lead to the accumulation of immune cells and pro-inflammatory cytokines within the perivascular compartment, resulting in PVS dilation and further compromise of interstitial fluid drainage.^61^ This cascade could contribute to the accumulation of neurotoxic metabolites and sustained neuroinflammation, creating a self-perpetuating cycle that manifests clinically as persistent cognitive dysfunction.^62-64^ The observation that PVS-cognition associations were most robust for measures of processing speed and executive function—capacities that are highly dependent on white matter integrity and the efficiency of distributed neural networks—is consistent with this model. These findings align with recent studies demonstrating glymphatic dysfunction in neuroPASC participants using diffusion-based metrics and provide complementary evidence from a structural MRI perspective. ^26,65^

Several limitations should be considered in interpreting these results. First, the cross-sectional design precludes causal inference regarding the directionality of PVS-cognition associations. Longitudinal studies are needed to determine whether PVS burden precedes, accompanies, or follows cognitive decline in neuroPASC. Second, the sample size, while adequate for detecting moderate effect sizes, limited statistical power for some subgroup analyses, particularly among participants without fatigue (n = 5) and without brain fog (n = 8), which precluded analysis in that subgroup. Third, the recruitment criteria evolved over the study period, initially based on self-reported neurological symptoms and later incorporating the neuroPASC diagnostic classification after the classification criteria were formally established in 2023, which may introduce some heterogeneity.^66^ Fourth, PVS quantification was performed using a semi-automated tool (PVSSAS) that, while validated, involves some degree of operator judgment. Finally, the neuropsychological battery, while including well-validated measures, did not comprehensively assess all cognitive domains that may be affected in neuroPASC.

Despite these limitations, the use of 7T MRI represents a significant methodological strength, enabling the detection and quantification of PVS with resolution and contrast that would not be achievable at conventional clinical field strengths. The inclusion of both lobe-specific PVS analyses and symptom-defined subgroup analyses provides a more nuanced understanding of brain-behavior relationships in this population than has previously been reported.

## Conclusion

This study demonstrates that while global PVS burden does not differ between neuroPASC participants and comparators, PVS metrics are significantly associated with cognitive performance among neuroPASC participants, particularly in the domains of processing speed and executive function. These brain-behavior associations were most pronounced in frontal regions. These findings suggest that PVS burden, as a potential marker of impaired glymphatic function and neuroinflammation, may contribute to the pathophysiology of cognitive dysfunction in neuroPASC. Future longitudinal studies should investigate the trajectory of PVS changes over time in neuroPASC and determine whether PVS burden can serve as a prognostic biomarker or therapeutic target for cognitive rehabilitation in this population.

## Data availability

The data that support the findings of this study are available from the corresponding author, upon reasonable request.

## Acknowledgements

The authors gratefully acknowledge Dr. David Putrino and Dr. Benjamin Natelson for their valuable assistance with participant recruitment for this study.

